# TransferGWAS of T1-weighted Brain MRI Data from the UK Biobank

**DOI:** 10.1101/2024.06.11.24308721

**Authors:** Alexander Rakowski, Remo Monti, Christoph Lippert

## Abstract

Genome-wide association studies (GWAS) traditionally analyze single traits, e.g., disease diagnoses or biomarkers. Nowadays, large-scale cohorts such as the UK Biobank (UKB) collect imaging data with sample sizes large enough to perform genetic association testing. Typical approaches to GWAS on high-dimensional modalities extract predefined features from the data, e.g., volumes of regions of interest. This limits the scope of such studies to predefined traits and can ignore novel patterns present in the data. TransferGWAS employs deep neural networks (DNNs) to extract low-dimensional representations of imaging data for GWAS, eliminating the need for predefined biomarkers. Here, we apply transferGWAS on brain MRI data from the UKB. We encoded 36, 311 T1-weighted brain magnetic resonance imaging (MRI) scans using DNN models trained on MRI scans from the Alzheimer’s Disease Neuroimaging Initiative, and on natural images from the ImageNet dataset, and performed a multivariate GWAS on the resulting features. Furthermore, we fitted polygenic scores (PGS) of the deep features and computed genetic correlations between them and a range of selected phenotypes. We identified 289 independent loci, associated mostly with bone density, brain, or cardiovascular traits, and 14 regions having no previously reported associations. We evaluated the PGS in a multi-PGS setting, improving predictions of several traits. By examining clusters of genetic correlations, we found novel links between diffusion MRI traits and type 2 diabetes.

**Author Summary:** Genome-wide association studies are a popular framework for identifying regions in the genome influencing a trait of interest. At the same time, the growing sample sizes of medical imaging datasets allow for their incorporation into such studies. However, due to high dimensionalities of imaging modalities, association testing cannot be performed directly on the raw data. Instead, one would extract a set of measurements from the images, typically using predefined algorithms, which has several drawbacks - it requires specialized software, which might not be available for new or less popular modalities, and can ignore features in the data, if they have not yet been defined. An alternative approach is to extract the features using pretrained deep neural network models, which are well suited for complex high-dimensional data and have the potential to uncover patterns not easily discoverable by manual human analysis. Here, we extracted deep feature representations of brain MRI scans from the UK Biobank, and performed a genome-wide association study on them. Besides identifying genetic regions with previously reported associations with brain phenotypes, we found novel regions, as well as ones related to several other traits such as bone mineral density or cardiovascular traits.

## 2 Introduction

The growing size of medical imaging datasets within biobanks is increasing the power of genome-wide association studies (GWAS) performed on such modalities. For example, the number of associated loci found in a GWAS of phenotypes derived from brain magnetic resonance imaging (MRI) data in the UK Biobank (UKB) increased over 4-fold between the initial study of Elliott et al. [14] and the consecutive study of Smith et al. [49]. The initial approaches to imaging GWAS were based on the extraction of predefined image-derived phenotypes (IDPs) [14, 49, 43]. While being interpretable, such analyses require the availability of automated tools for IDP extraction for the modality of interest and are limited to traits defined a priori, potentially preventing novel genetically-driven phenotypes from being discovered.

Instead of using manually defined traits, a recent line of work employed deep learning (DL) to derive imaging features using pretrained deep neural network (DNN) models to perform GWAS on. This approach has been demonstrated to be successful in a range of imaging modalities, including retinal fundus images [24, 53], cardiovascular magnetic resonance (CMR) images [3, 4], or brain MRI scans [39]. In this work, we perform an imaging GWAS on *N* = 36, 311 T1-weighted brain MRI scans from the UKB dataset. As opposed to the ENDO approach of Patel et al. [39], who pretrained a DNN on data from the same dataset where the GWAS was performed, we employed the transferGWAS pipeline of Kirchler et al. [24] and used two DNN models pretrained on other datasets, following a transfer learning methodology. We encoded the brain scans using models pretrained on the ImageNet [48] and Alzheimer’s Disease Neuroimaging Initiative (ADNI) datasets [35], with the former extracting “general” image features and the latter focusing on brain MRI and dementia-specific ones. Our GWAS performed on these features identified a number of loci not detected in the IDP or ENDO brain MRI studies, several of which were not reported in any previous GWA studies. We further conducted downstream analyses using the discovered genetic variants, demonstrating their utility in creating more predictive polygenic score (PGS), and pointing to novel genetic correlations between type 2 diabetes (T2D) and diffusion magnetic resonance imaging (dMRI) traits (see Fig 1 for an overview of our workflow).

**Figure 1:**
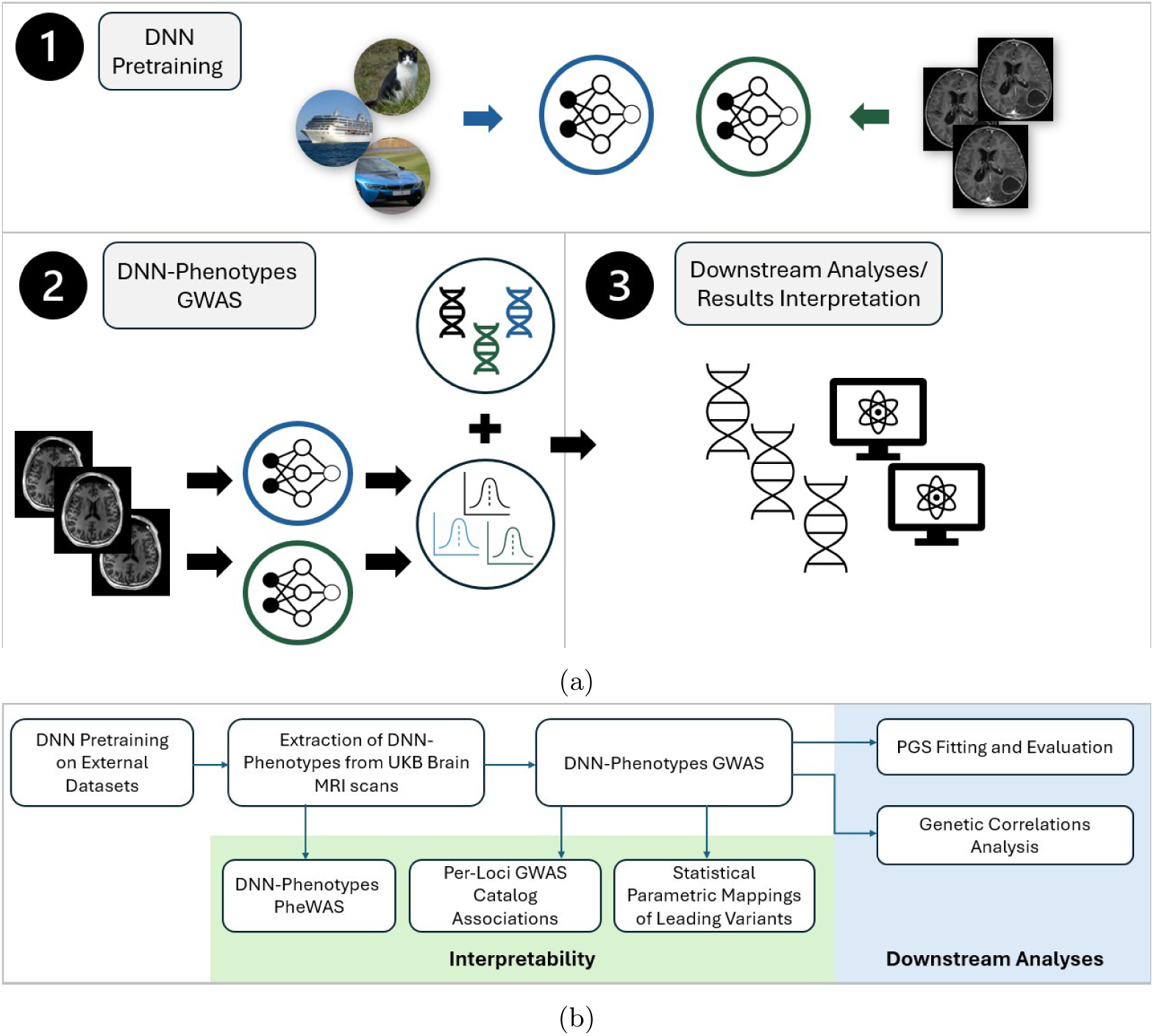
Overview of our study and workflow. **(a)** A general overview of the study: (1) - we pretrained 2 DNN models on external datasets of natural images, and of brain MRI scans (2) - encoded brain MRI data from the target datasets and performed GWAS on the DNN-derived phenotypes (3) performed a series of downstream analyses using the learned DNN features and discovered genetic variants. **(b)** Description of each step involved in the complete workflow of our study.

## 3 Results

### 3.1 Interpretation of the DNN Features

In order to interpret the signal carried by the DNN features, we extracted the first 10 principal components (PCs) of both DNN models, and performed a phenome-wide association study (PheWAS) against each PC and 7, 744 UKB phenotypes (supplementary Table S2). We found 2, 408 and 2, 622 significantly associated phenotypes for the ImageNet and ADNI PCs respectively, having P-values below the Bonferroni-corrected threshold of *≈* 6.5 *·* 10*^−^*^7^. Fig 2 shows the percentage of significantly associated traits per category. The top 35 categories with the highest ratio of significant hits contained 17 brain-related categories, with the other ones being bone density, body composition, or blood-related categories. In almost all cases the ADNI PCs were associated with a higher number of phenotypes than the ImageNet PCs.

**Figure 2:**
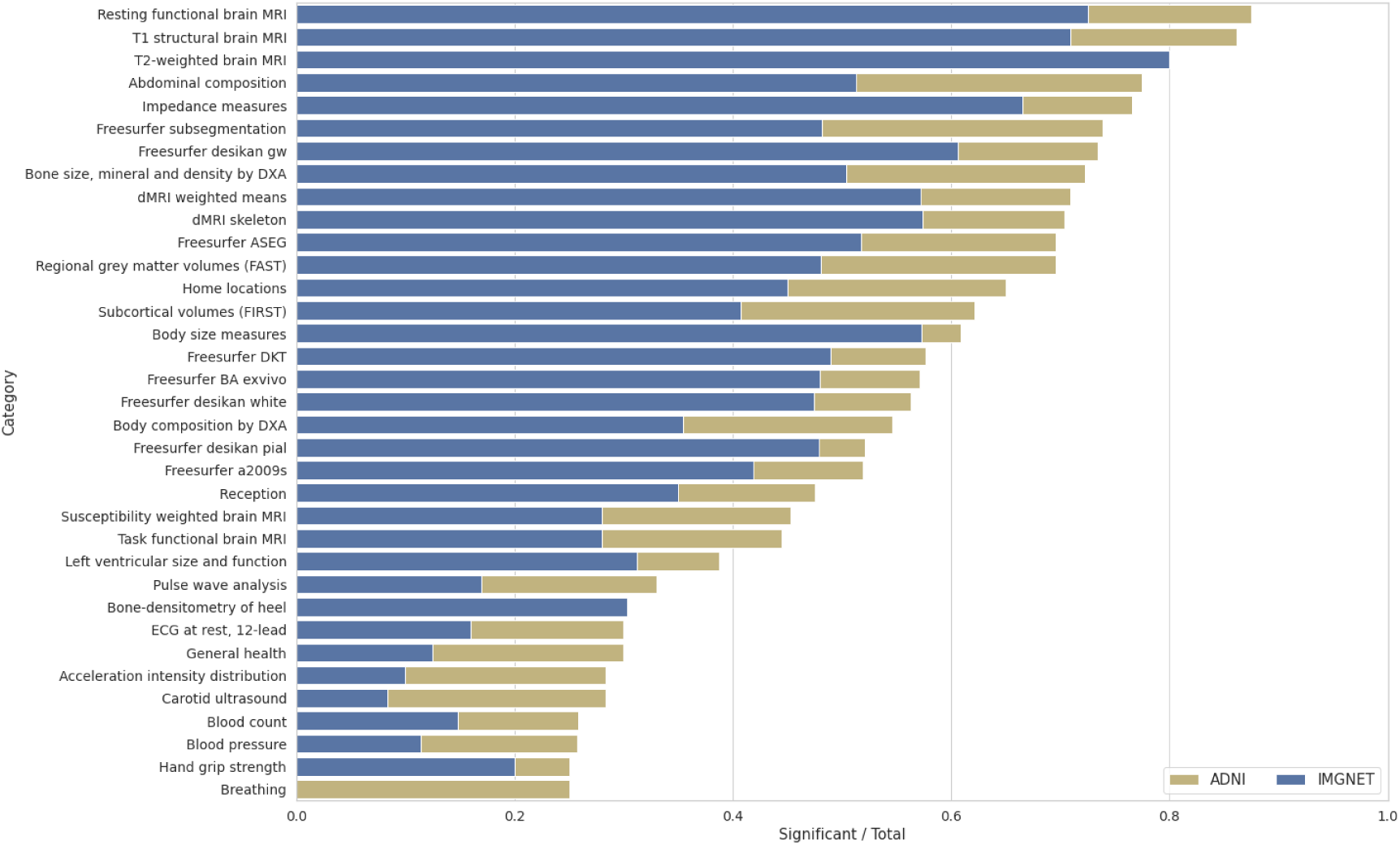
Results of the PheWAS performed on the principal components (PCs) of the ImageNet (blue) and ADNI (yellow) pretrained models. For each phenotype category from the UK Biobank (UKB) we plot the number of significant associations per model divided by the total number of traits in that category. Shown are the top 35 phenotype categories with the highest ratio of significant associations.

### 3.2 GWAS Results

At the Bonferroni-corrected significance threshold of 2.5 *·* 10*^−^*^9^, we found 4, 665 peak associations for the ImageNet and 5, 291 for the ADNI pretrained models, resulting in 4, 382 and 4, 360 distinct variants for ImageNet and ADNI. The clumping procedure then identified 194 and 165 independent regions for the ImageNet and the ADNI models respectively. This amounted to 7, 458 distinct variants and 289 distinct regions across all 20 features of both DNN models. Fig 3 shows the Manhattan plots for both models, aggregated over each of the 10 PCs per model. We estimated the heritability of each PC using linkage disequilibrium score regression (LDSC) (Section 5.5) and found the ADNI-pretrained PCs to be more heritable, with a mean *h*^2^ = 0.19, and the ImageNet PCs having a mean *h*^2^ = 0.13 (Fig 4). The summary statistics for all PCs are made publicly available as a figshare resource under https://doi.org/10.6084/m9.figshare.25933717.v1.

**Figure 3:**
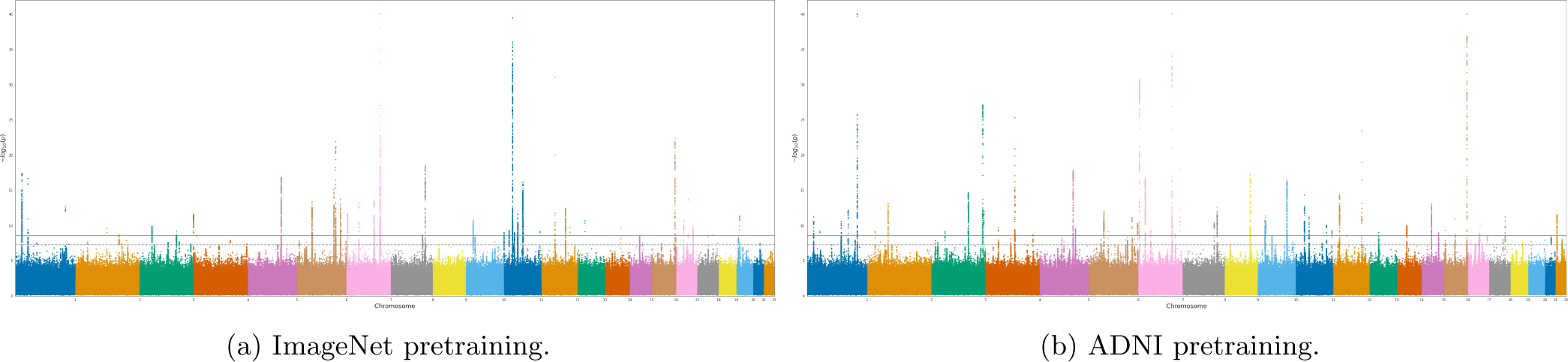
Manhattan plots for the GWAS (*n* = 36, 311 individuals, 16, 472, 121 SNPs) performed on features of the ImageNet (plot *a*)) and the ADNI (plot *b*)) pretrained models. The horizontal lines mark the initial significance threshold of 5 *·* 10*^−^*^8^ (dashed line) and Bonferroni-corrected threshold of 2.5 *·* 10*^−^*^9^ (solid line). For visualization purposes we truncate P-values below 10*^−^*^40^ and plot only the minimal P-values across each of the 10 features per model.

**Figure 4:**
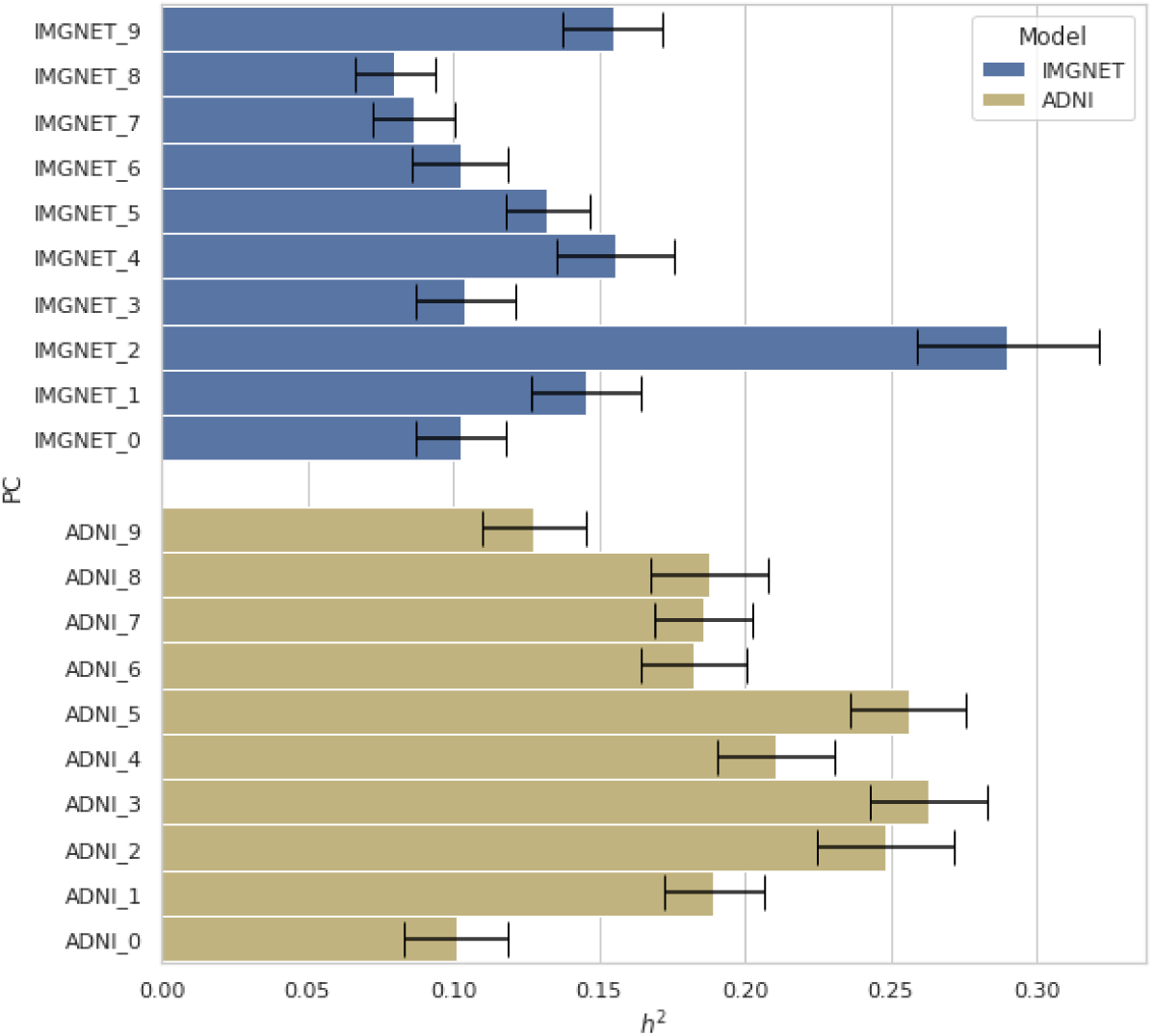
*h*^2^ heritability estimates of principal components (PCs) of the ImageNet (blue) and ADNI (yellow) pretrained models, obtained using linkage disequilibrium score regression (LDSC). Black lines indicate the standard error of the estimates.

#### 3.2.1 GWAS Catalog Associations

For each independent locus, we queried associations reported in previous GWA studies from the NHGRI-EBI GWAS Catalog [7] (Fig 5). The dominating phenotype categories included bone mineral density (BMD)-related traits and a range of brain traits, such as cortical thickness, diffusion, or volumes of brain regions of interest (ROIs). We note that the ADNI-pretrained features tagged more regions corresponding to brain-related traits, whereas the ImageNet model tagged more regions related to “general” body structure, such as BMD, height, or body mass index (BMI). Overall, out of the 289 independent loci, 72 did not have brain-related associations reported in the catalog.

**Figure 5:**
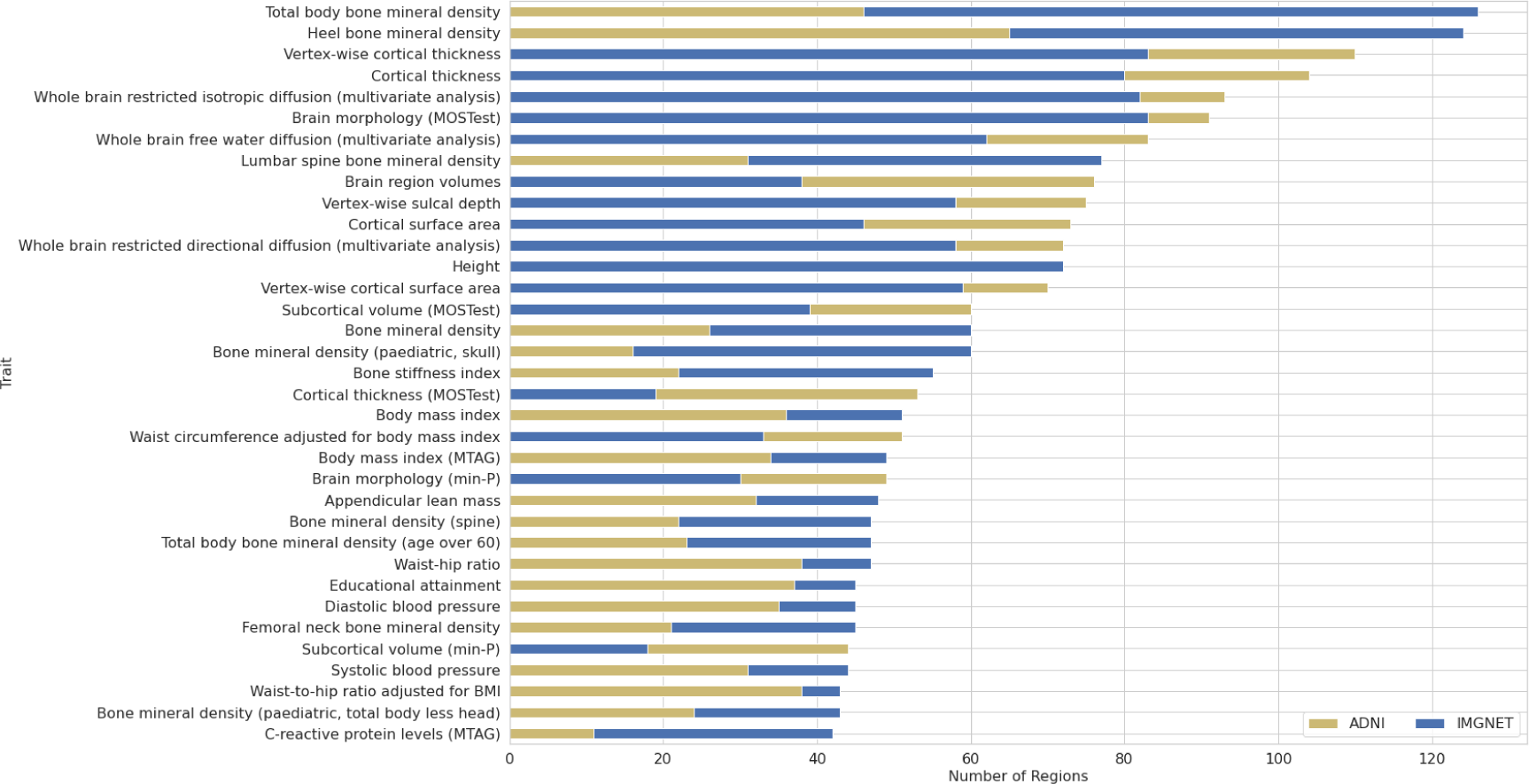
Number of independent loci per trait with associations reported in previous studies included in the NHGRI-EBI GWAS Catalog [7]. Shown are the top 35 traits with the highest number of associated regions.

Among neuropsychiatric disorders with the highest number of distinct regions, 47 were associated with schizophrenia, 37 with neuroticism, 36 with attention deficit hyperactivity disorder, 35 with bipolar disorder, 33 with depression, 32 with Alzheimer’s disease, 30 with autism, 22 with anorexia nervosa and 21 with anxiety.

3 out of the 10 first traits were not directly brain-related: heel bone mineral density (HBMD) (144 regions), total BMD (125 regions), and height (113 regions). The associations between BMD and the brain have been investigated in the context of neurological disorders [29, 30, 54], as well as in samples of healthy subjects [2]. Loskutova et al. [29, 30] reported a correlation between BMD and an early onset of Alzheimer’s disease (AD), as well as with several brain volumes. HBMD is postulated to be a causal factor for multiple sclerosis (MS) through an increased risk of fractures [54]. Bae et al. [2] showed that osteoporosis increases the pace of parenchymal atrophy and ventricular enlargement during aging of healthy individuals.

Another prevalent category were blood-related traits, such as cell counts: white (67), red (32), monocyte (45), neutrophil (41), eosinophil (40), lymphocyte, (25) reticulocyte (23), blood pressure (95) or hypertension (45), or hemoglobin (68). Blood pressure and hypertension are known factors influencing brain morphology, as well as cognitive performance or dementia [50, 13, 15, 45], while aenemia is a causal factor for cognitive decline and AD [40, 52].

#### 3.2.2 Novel Loci

In total, we found existing associations for 275 regions in studies conducted on the British population, and 278 regions among all populations. Out of the remaining 11 loci, one was located within an RNA gene, and 10 within 7 distinct protein-coding genes. Among the associated phenotypes, 6 genes were associated with mental or neurodevelopmental disorders such as AD, schizophrenia, or attention-deficit/hyperactivity disorder (ADHD), and 4 genes were associated with T2D.

As a further means of interpreting the novel regions, we computed statistical parametric mappings (SPMs) for each leading single-nucleotide polymorphism (SNP) (Fig 6) and computed the fractions of volume of each brain region correlated with each lead variant (Fig 7). All SNPs were correlated with the left cerebral cortex and cerebrospinal fluid. Most notable was the variant rs111469125 (16:87268090) located inside the C16orf95 gene, being correlated with 19 out of 22 brain regions, in particular with several ventricle structures: the 3rd and 4th ventricles (3% of total voxels), and the left and right lateral ventricles (1% and 1.5% of total voxels). It was also correlated with 3% of the voxels of cerebrospinal fluid, and was the only new variant correlated with the left cerebellum white matter, the left thalamus, and the right caudate.

**Figure 6:**
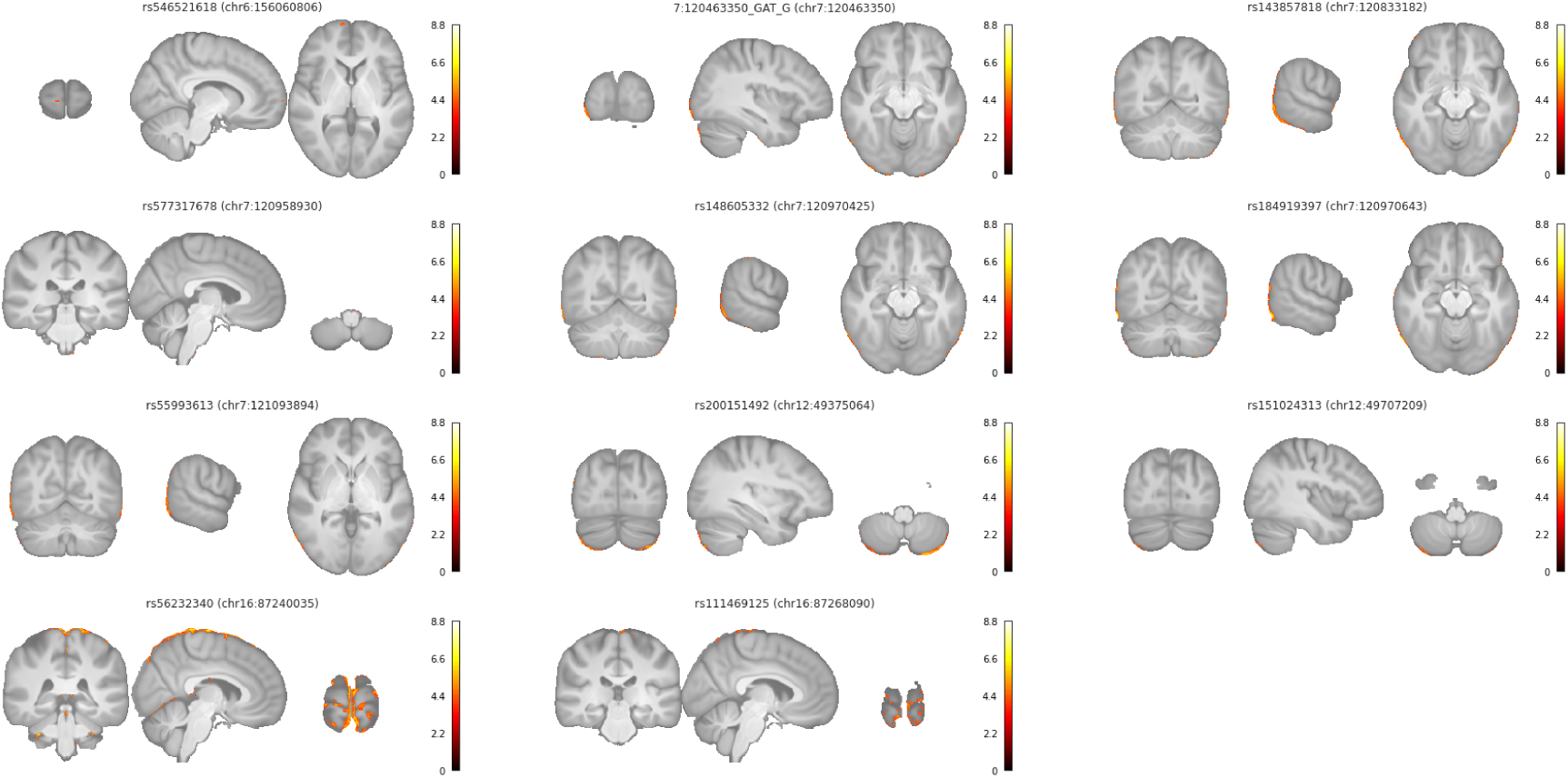
Statistical parametric mappings (SPMs) for genetic regions with no previously reported GWAS associations. Plotted are values of the *t* -statistics of the correlation coefficients between lead variants of each region and each single voxel in the MRI scans. We plot values below the Bonferroni-corrected significance threshold accounting for the total number of voxels tested.

**Figure 7:**
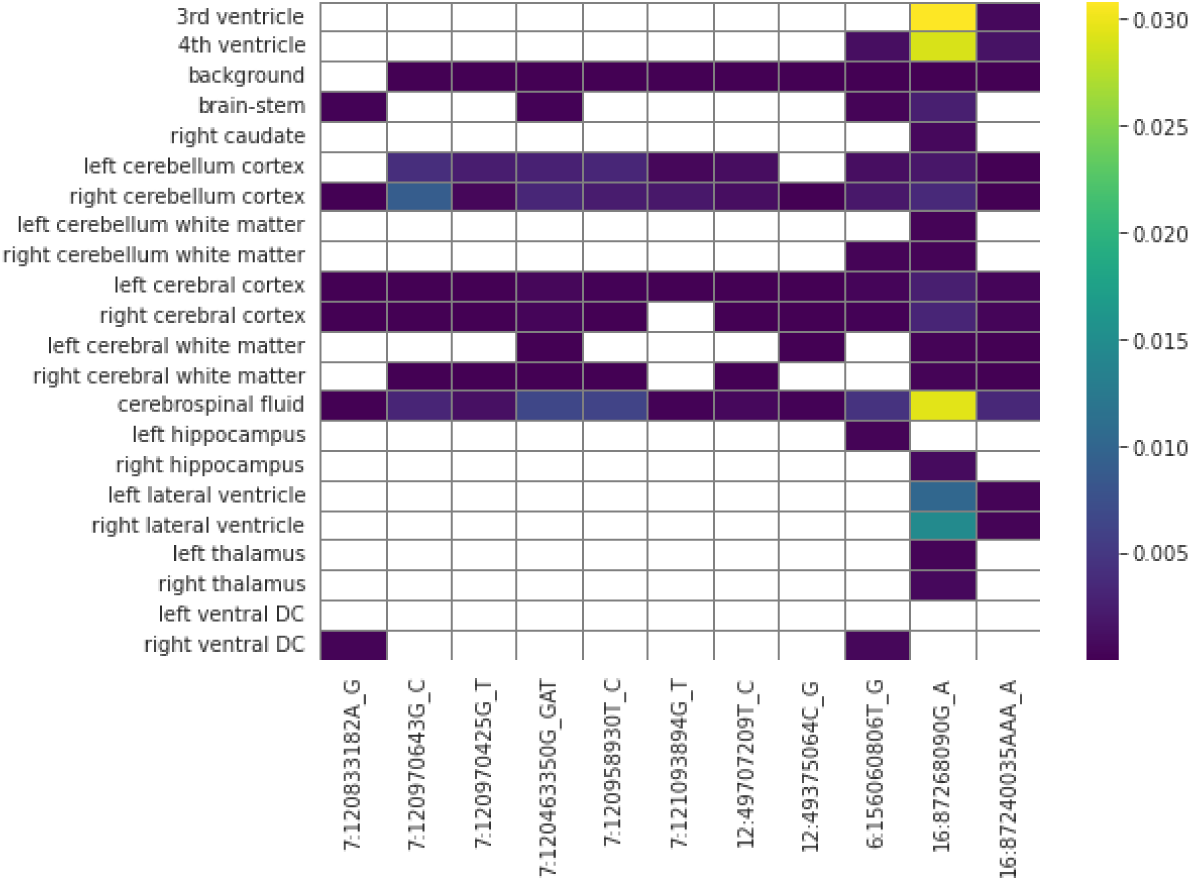
Fractions of volume of brain regions correlated with genetic regions with no previously reported GWAS associations. The values are computed as the total number of voxels in a given brain region significantly correlated with a lead variant, divided by the total number of voxels in that brain region. White cells indicate no voxels being significantly correlated for a given brain region-genetic region pair.

#### 3.2.3 Comparison with Previous Studies

We performed another GWAS using discovery and replication cohorts (23,604 and 12,709 samples), replicating 1,631 hits over 1,510 unique variants, which amounted to 70 replicated loci. We compared our results with two GWA studies on UKB brain MRI data - the first one using 3,144 brain imaging-derived phenotypes [49] and the second study using 256 DL-based features [39], which yielded 692 and 43 replicated loci respectively. Out of our 70 replicated loci, 9 were not present in the 692 of Smith et al. [49], and 28 were not present in the 43 loci of Patel et al. [39].

### 3.3 TransferGWAS Polygenic Scores

Here, we evaluated the potential of variants discovered in our study for downstream prediction of phenotypes using 20 PGS fitted for each of the 20 DNN PCs with the summary statistics from our GWAS. In order to compute the features of the DNN models, imaging data need to be present, which constitutes less than a tenth of all UKB samples. On the other hand, genotyping data were available for all participants. This allowed us to calculate the PGS for all remaining *N* = 451, 450 participants not included in the GWAS sample. The corresponding methods are described in Section 5.4, while the weights of the fitted scores are made publicly available as a figshare resource under https://doi.org/10.6084/m9.figshare.25933663.v1.

#### 3.3.1 PGS PheWAS

To gain insights into which traits the PGS might be predictive of, we performed a PheWAS on the 20 PGS and the 7, 744 UKB phenotypes (supplementary Table S2). Note that while the “raw” DNN PCs can encode both genetic and environmental signals, the PGS should capture only the former, and thus we expected the associations between the phenotypes to differ from the PheWAS performed on the PCs. The total number of significant PC-phenotype associations and the effect sizes were higher for the original PCs than for the PGS: 28, 767 vs. 25, 948 significant associations in total, 2, 928 vs. 2, 860 distinct associated traits, with mean effect sizes of 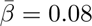 vs. 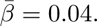 We identified 3 potentially interesting groups of associations (Fig 8):

- traits related to BMD
- weight/fat mass/BMI
- cardiovascular traits

**Figure 8:**
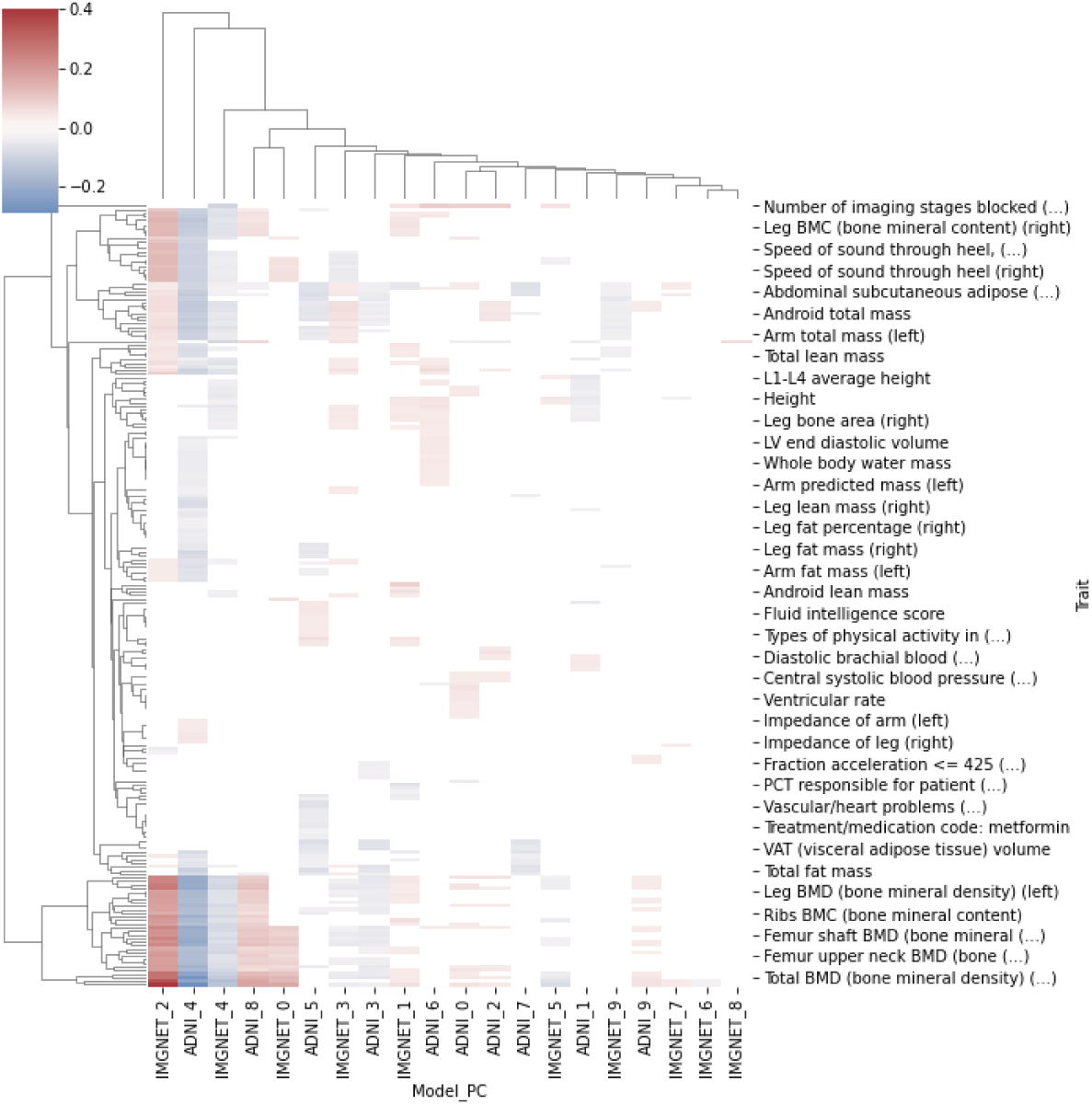
Results of the PheWAS (*N* = 451, 450) between the polygenic scores (PGS) fitted on the features of the DL models (rows) and phenotypes from the UK Biobank (UKB) dataset (columns). Cell colors represent the magnitudes and the signs of the estimated association coefficients between each PGS and phenotype combination.

which we decided to investigate further in a prediction setting.

#### 3.3.2 Predictive Performance Compared to Trait-Specific PGS

We tested the utility of our developed PGS by evaluating whether they can improve predictions of phenotypes from UKB over PGS designed specifically for particular traits in a multi-PGS setting [25]. We chose a set of 9 phenotypes based on the PheWAS results and computed their corresponding scores using PGS available in the PGS Catalog [26]. For each phenotype, we then fitted and evaluated two linear models: one fitted using only the trait-specific PGS, and one additionally using our transferGWAS PGS. While there were statistically significant improvements in predictions for 4 out of 9 traits, they yielded arguably small performance increases (*∼* 1.5% of relative improvement), with the exception of predicting HBMD using a (general) BMD PGS, where the relative improvement was over 20% (Table 1). We decided to further investigate the HBMD results. Since the improvement could have been stemming from a lower signal in the dataset of the external PGS, compared to the UKB, we conducted a further comparison within the UKB. We performed a GWAS on HBMD using 19, 909 samples from our GWAS data which had HBMD measurements available, and 16, 404 randomly drawn from the remaining samples of the white-british participants, to match the sample size and population of our GWAS, and fitted a new PGS on the resulting summary statistics. We then evaluated the newly-created HBMD PGS with and without our transferGWAS PGS on the remaining UKB data and observed the same relative improvement of 1% in performance (*p <* 0.001). This indicates that transferGWAS has the potential to identify additional variants for related traits while using the sample size. We hypothesize that this might be due to certain pleiotropic variants having a larger effect on the DNN PCs than on HBMD, and thus being able to be detected with our DL GWAS and not with the HBMD-dedicated GWAS.

**Table 1:** Comparison of predictive performance of Multi-PGS models using only trait-specific polygenic scores (PGS) (2nd column) and including our TransferGWAS PGS (3rd column) for a set of selected phenotypes from UK Biobank (UKB), measured with the *R*^2^ coefficient of determination. Significant differences are marked with (*). Heel bone mineral density (1) and (2) correspond to results of using PGS for heel bone mineral density, or (general) bone mineral density respectively.

| Trait | PGS Catalog | Our PGS + PGS Catalog | $\Delta$ |
| --- | --- | --- | --- |
| Height | 0.759 | 0.759 | 0.000 |
| BMI | 0.284 | 0.284 | 0.000 |
| Heel bone mineral density (1) | 0.273 | 0.275 | 0.002* |
| Heel bone mineral density (2) | 0.064 | 0.078 | 0.014* |
| Red blood cell count | 0.387 | 0.387 | 0.000 |
| White blood cell count | 0.104 | 0.106 | 0.002* |
| Systolic blood pressure | 0.283 | 0.284 | 0.000 |
| Diastolic blood pressure | 0.175 | 0.176 | 0.001* |
| Ventricular rate | 0.027 | 0.025 | -0.001 |

### 3.4 Genetic Correlations

The results of the PheWAS conducted on the learned PCs led us to a set of traits that we decided to investigate further. In order to analyze the genetic components of the PCs, we computed genetic correlation coefficients between 102 selected traits and each of the 20 PCs (see Section 5.5 for details). 39 traits were significantly correlated, surpassing the Bonferroni-corrected threshold of *≈* 2.5 *·* 10*^−^*^5^. We grouped the traits into 3 groups:

- (volumes of) brain ROIs (e.g., ventricles, brain stem, cerebrospinal fluid (CSF))
- dMRI traits (e.g., fractional anisotropy (FA), orientation dispersion index (ODI))
- “general” traits: Height, T2D, BMI, HBMD

Additionally, we tested for correlations with AD, educational attainment, and unipolar depression, finding no significant correlations when corrected for multiple testing (*p >* 0.001). The significantly associated traits are shown in Fig 9, where we observed several “clusters” of PC-trait associations.

**Figure 9:**
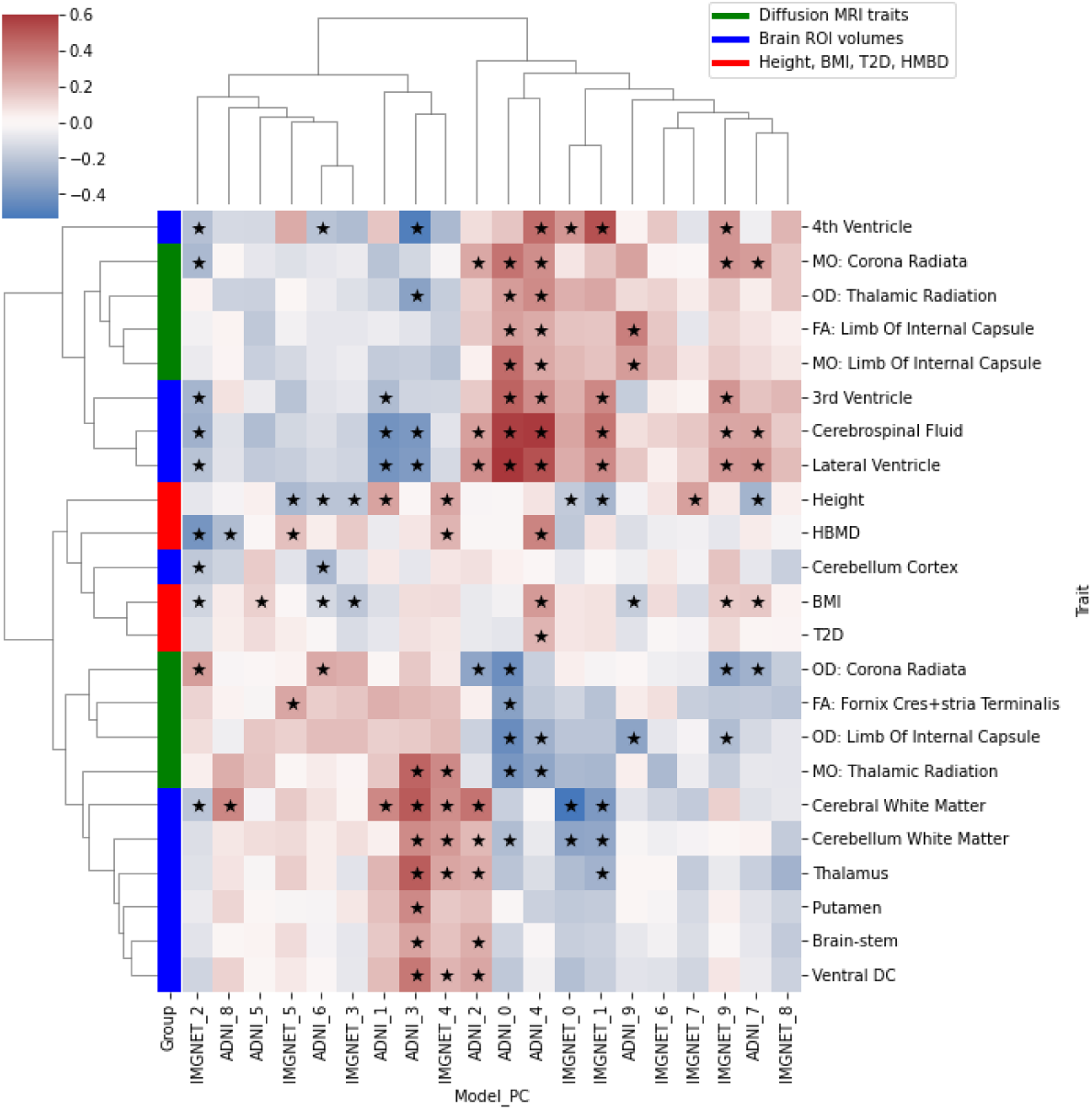
Genetic correlation coefficients between the 20 deep neural network (DNN) principal components (PCs) (rows) and 23 significantly associated phenotypes (columns), out of 27 candidate traits from the UK Biobank (UKB). Cell colors represent the magnitudes and the signs of the estimated genetic correlation coefficients between each PC and phenotype combination.

Several PCs were associated with volumes of multiple brain ROIs. The first two PCs of ImageNet (IMGNET0, IMGNET1) seemed to capture the overall body size, as they were negatively correlated with height and white matter volume, and positively with ventricular ROIs and CSF.

PCs ADNI2, ADNI3, and IMGNET4 were genetically associated with volumes of several brain ROIs, e.g., cerebral white matter, putamen, or thalamus. ADNI2 and ADNI3 were also associated with volumes of CSF and the lateral ventricle. Interestingly, ADNI2 had a positive correlation both for CSF and the lateral ventricle, as well as for gray and white matter structures, whereas one might expect the ventricular volumes (and thus CSF) to grow with the shrinkage of brain structures.

PCs associated with HBMD seemed to capture different aspects of brain anatomy. IMGNET2 had a negative correlation with HBMD, BMI, and cerebral white matter, but also with multiple ventricular volumes. On the other hand, ADNI8 and IMGNET4 also had negative genetic correlations with HBMD, but positive ones with cerebral white matter.

ADNI0 and ADNI4 were associated with a range of Diffusion MRI traits, as well as with several ventricular ROIs. Furthermore, ADNI4 was genetically correlated with HBMD and BMI, and was the only PC associated with T2D, which we further discuss below.

#### 3.4.1 ADNI4 and T2D

BMI was shown to increase the risk of developing T2D [8, 17], as well as being genetically correlated to T2D [8]. The signs of genetic correlations between ADNI4, and BMI and T2D were also matching. ADNI4 was also positively genetically associated with HBMD. T2D patients have been shown to have a higher bone density [42, 27]. Evidence also exists for shared heritability between BMD and T2D, albeit relatively small [47, 55]. As with BMI, the sign of the genetic correlation between ADNI4 and HBMD was positive. Regarding the brain ROIs, ADNI4 was positively correlated with volumes of the lateral, 3rd, and 4th ventricles, as well as with the CSF. Ventricular enlargement and increase in CSF are associated with several neurodegenerative diseases, such as AD, MS, or schizophrenia [9, 36]. Several studies showed an association between T2D and volumes of white matter structures (whole brain volume, frontal lobe), gray matter (overall trend in all structures), as well as CSF and ventricular volumes [33]. Furthermore, ADNI4 was genetically correlated with 35 different dMRI traits:

##### Mean diffusivity (MD) traits

4 MD traits were positively genetically correlated with ADNI4: fornix, superior serebellar peduncle (both sides), and the superior fronto-occipital fasciculus (left). Positive associations between T2D and MD have been found in observational studies [19, 46].

##### Fractional anisotropy (FA) traits

FA traits have been found to be negatively correlated with T2D in literature [19, 46, 33]. We found 4 traits to be negatively genetically correlated with ADNI4, however the posterior limb of left internal capsule was positively genetically correlated with the PC. The direction of this correlation seemed to be in opposition to the associations found in observational studies [33]. On the other hand, it is postulated to be causal with the same sign for fasting insulin [12], an increase of which is an indicator of T2D. We identified two regions containing shared variants located at Chr2:27766284 and Chr14:91881387. The first region contains missense and intron variants for GCKR gene (ENSG00000133962), a glucokinase regulator, with no previously reported associations for brain phenotypes, missense and intron variants for C2orf16 (ENSG00000221843) and intron variants for ZNF512 (ENSG00000243943) both protein coding genes with association with neurodegenerative diseases, T2D, and blood measurements. The second region contains intron variants for the CCDC88C (ENSG00000015133), a protein coding gene, with associations with glucose metabolism, brain measurements, and neurodegenerative diseases, and CCDC88C-DT (ENSG00000258798), and RNA gene that is a divergent transcript for CCDC88C, with associations with brain measurements and hypertension. The above may be another indicator of a non-trivial relation between FA of limb of internal capsule and T2D, with potentially different shared heritability and environmental effects.

##### Orientation dispersion index (ODI) traits

7 ODI traits were positively correlated with ADNI4, while 3 traits were correlated negatively. ODI of white matter tracts was reported to be positively correlated with duration of T2D and with levels of HbA1c, a marker for T2D, while ODI of internal capsule was reported to have a negative correlation [1], which is consistent with 9 out of 10 of our findings. We found a negative genetic correlation for the posterior right corona radiata, which had shared variants in regions located at chr8:119486034 and 11:27465591. The first region has intron variants for SAMD12 (ENSG00000177570), a protein coding gene with associations with brain measurements, MS, bone density, and blood measurements. The second region has intron variants for LGR4 (ENSG00000205213), a protein coding gene with associations with brain measurements, bone density, and body mass traits.

##### Mode of anisotropy (MO) traits

5 MO traits were genetically positively correlated with ADNI4, and 6 negatively. Fasting insulin, a marker for T2D was reported to be negatively associated with anterior corona radiata [12]. We found positive genetic correlations between both sides of the posterior and superior corona radiata and ADNI4, with shared variants with T2D located in the region chr2:27766284 for the superior, and in chr8:119486034 for the posterior. The first region contains missense and intron variants for the GCKR and C2orf16 genes, and an intron variant for ZNF512 (see the FA regions), and the second region has intron variants for SAMD12 (see the ODI regions above). The correlations between the other 7 traits are reported in supplementary Table S3.

## 4 Discussion

Using the transferGWAS approach we performed a GWAS on 20 DNN feature representations of 36, 311 T1-weighted brain MRI scans from the UKB, identifying 289 loci, 11 of them without any previously reported associations, and 72 without any associations for brain-related traits. Similar to the findings of the initial transferGWAS study of retinal fundus images of Kirchler et al. [24], the features of an ImageNet-pretrained model were associated with a higher number of loci related to “general” body structure traits, such as BMD or BMI, whereas features from a model pretrained directly on brain MRI data identified more loci corresponding to brain measurements and neurodegenerative diseases. Overall, features of both DNN models were associated directly, through PheWAS, or genetically, through GWAS-identified loci, with a large number of BMD traits. For example, the ImageNet and ADNI-derived features were significantly associated with over 50% and 70% of phenotypes under the UKB category 125 “Bone size, mineral, and density by DXA”, and with over 120 and 40 distinct loci associated “Total body bone mineral density” in the NHGRI-EBI GWAS Catalog. Detecting these genetic regions in features derived from brain MRI data seems to confirm the connections between BMD and brain measurements, as well as with neurodegenerative diseases previously reported in the literature (as discussed in Section 3.2.1), which we further investigated with an analysis of genetic correlations (Section 3.4), highlighting particular brain ROIs genetically associated with BMD. Furthermore, the genetic correlations identified by our study shed more light on the relations between dMRI measurements and T2D, BMI, as well as cardiovascular traits, also reported in several studies (Section 3.4.1). Finally, we demonstrated a practical application of our findings by constructing PGS of our DNN-derived phenotypes, which improved predictions of existing PGS of BMD, white cell blood count, or diastolic blood pressure. In a further analysis, we fitted a PGS directly to HBMD measurements on a UKB sample of the same size as our GWAS and observed the same improvement in performance when augmented with our DNN PGS, indicating that the transferGWAS approach can identify additional variants for a trait of interest, being complementary to conducting a trait-dedicated GWAS.

We demonstrated how transferGWAS can be applied to discover new variants and in turn, lead to better phenotype predictions. However, a drawback of using features of pretrained DNN models as traits of interest is their reduced interpretability compared to predefined phenotypes. While we analyzed both the DNN-derived traits and the discovered loci with a range of techniques (PheWAS, querying the GWAS Catalog, SPMs), we highlight the need for further developing apossibly automated pipeline for interpretability of the DNN features, to foster their utility for consecutive research and clinical applications.

## 5 Materials and Methods

### 5.1 Pretraining of the Neural Network Models

The first model used for feature extraction was trained on 4,480 T1-weighted scans from the ADNI dataset [41]. The network architecture was a 3D convolutional variational autoencoder (VAE) [23], trained in a multi-task manner. The model consisted of 3 sub-networks: an encoder, a decoder, and a prediction head. The 256-dimensional outputs of the encoder network constituted the latent representations of the input data. The first task was the standard VAE objective, i.e., reconstructing the input scans from the latent representations, while regularizing the representations to match a standard normal prior distribution with a Kullback-Leibler divergence (KLD) loss term. The second task was to predict the clinical dementia rating (CDR) from the latent representations. The aim of the VAE objective was to learn general structural features describing an MRI scan, while the prediction task should promote neurodegenerative features associated with the presence of dementia. Additionally, we input the age and sex of each participant into the decoder and prediction networks, forcing the model to learn latent representations invariant to age and sex, and thus potentially increasing the statistical power of the GWAS. We trained the model for 500 epochs with the Adam optimizer [22], with a mini-batch size of 128. The weights of the reconstruction, KLD, and the predictions loss terms were 1, 10*^−^*^5^ and 10*^−^*^2^ respectively. For data preprocessing, we skull-stripped each scan using using the HD-BET tool [20], performed a non-linear registration to the MNI152 template with a 1mm^3^ resolution using the FLIRT and FNIRT commands from the FSL software [21], and finally downsampled the scans to a size of 96 *×* 96 *×* 96 voxels each.

Following Kirchler et al. [24], we also employed a 2D ResNet50 [18] model pretrained on ImageNet, a non-medical dataset of natural images [48]. We used a readily available trained model from the PyTorch library [38]. We selected the 2048-dimensional output of the penultimate layer as the latent features used for the GWAS. Since the model was trained on 2D data, we could not directly extract features from the 3D MRI scans. Instead, for each scan, we computed the features over each single slice across the axial axis and averaged the results into a single vector.

### 5.2 GWAS

We selected a sample of *N* = 36, 311 UKB participants who “self-identified as ‘White British’ and have very similar genetic ancestry based on a principal components analysis of the genotypes” (UKB field 22006). We performed the association testing within the linear mixed model (LMM) framework using the BOLT-LMM software [28]. We adjusted for confounding using age, sex, the identifiers of the genotyping array and UKB assessment center, and the first 10 genetic principal components. We filtered the SNPs with the following criteria: MAF*≥* 0.1%, Hardy-Weinberg Equilibrium with a significance level of 0.001, pairwise LD-pruning with *R*^2^ = 0.8, and maximum missingness of 10% per SNP and participant, which resulted in 577, 570 directly genotyped SNPs. Including imputed genotype data resulted in 16,472,121 variants in total, on which we performed the GWAS. We clumped the variants into independent loci using the PLINK software [44], with a physical distance threshold of 250kb and a significance threshold of 10*^−^*^9^ for the index SNPs. We queried the NHGRI-EBI GWAS Catalog [7] using the LDtrait web application [31], with an *R*^2^ cutoff of 10*^−^*^1^ and a 250kb window.

### 5.3 PheWAS

We performed the PheWAS on the PCs of both pretrained models using the PHESANT software [32], with a P-value threshold of *≈* 6.5 *·* 10*^−^*^7^ from the Bonferroni correction to account for 20 PCs and 7, 744 different phenotypes from UKB, adjusting for age and sex.

### 5.4 Polygenic Scores

We fitted the DNN PGS and the custom HBMD PGS using the PRScs method [16], with the prspipe software [37, 34]. For the predictive performance comparison, we queried the PGS Catalog [26] API for a list of PGS developed for each of the 9 phenotypes, ignoring scores that used the UKB for development, to avoid data leakage. We then computed scores for the *N* = 451, 450 participants who were not in our GWAS sample using the PGS Catalog Calculator [51]. For each phenotype, we fitted a baseline linear model using all corresponding trait-specific PGS and covariates (age, sex, UKB assessment center, UKB genotyping batch, all UKB genetic PCs) and another linear model which additionally included our 20 DNN PGS. We used 60% of the data for model fitting and evaluated it on the remaining 40%. We computed P-values for differences between achieved *R*^2^ scores of the two linear models using permutation tests with 1, 000 permutations, randomly selecting predictions from either model for each test sample in each permutation.

### 5.5 Genetic Correlations

To compute the genetic correlation scores between the PCs and selected traits, we used the LDSC method [6, 5]. We used the provided LD scores precomputed on 1000 Genomes data [10] over HapMap3 [11] SNPs, and used the default values for other parameters of the LDSC. In order to find regions potentially contributing to the genetic correlations between ADNI4, T2D, and dMRI traits (Section 3.4.1), we selected SNPs with a P-value below 0.0001 for which the magnitude of the product of the z-scores between both ADNI4 and T2D, and ADNI4 and a dMRI trait exceeded a threshold of 15. For the dMRI traits, we selected pairs where the sign of the product of the z-scores matched the sign of the genetic correlation with ADNI4. We consider a region a set of variants within 250, 000 base pairs from a “central” variant.

## Data Availability

Data produced are available as supplementary material (PheWAS summary statistics and genetic correlation results) and at figshare (GWAS summary statistics and polygenic scores weights)

https://doi.org/10.6084/m9.figshare.25933717.v1

https://doi.org/10.6084/m9.figshare.25933663.v1

https://github.com/HealthML/transfer-gwas-brain-mri

## 6 Acknowledgments

This research has received funding by the European Commission in the Horizon 2020 project INTERVENE (Grant agreement ID: 101016775), and was partially funded by the HPI Research School on Data Science and Engineering.

